# Expert in Ultrasound Skills: Feasibility of an IMU-video platform to describe technical profiles during focused cardiac ultrasound. Pilot study

**DOI:** 10.64898/2026.06.16.26355788

**Authors:** Felipe Riquelme, Sofia Rivera, Francisca Seydewitz, Barbara Lara, David Acuña

## Abstract

**Background:** Focused cardiac ultrasound (FoCUS) is operator dependent and requires coordinated probe manipulation, image interpretation and iterative visual feedback. Existing assessment approaches often emphasize final image quality or expert rating. We developed Expert in Ultrasound Skills (EXUS), a platform that synchronizes transducer-mounted inertial measurement unit (IMU) data with ultrasound video, and evaluated its technical feasibility during FoCUS acquisition.

**Methods:** This observational pilot study included 6 operators performing two repetitions of a four-view FoCUS protocol, yielding 12 analytical sessions and 48 planned acquisitions. Feasibility was defined by acquisition completion, video availability, start/stop events, fused IMU-video windows, temporal coverage, complete human label entries and IMU integrity. A 100-image Likert rating task was used to summarize pairwise inter-rater agreement for still-frame image quality assessment.

**Results:** All 48 planned acquisitions were completed with video, start/stop events, fused windows and complete human label entries. Temporal coverage was at least 90% in 47/48 acquisitions. IMU integrity endpoints exceeded the 80% threshold: 43/48 acquisitions had no extreme IMU-derived artifact, 43/48 had no active-segment IMU restart and 44/48 had no complete motion flatline. Mean pairwise exact agreement for the Likert task was 38.9%, with mean quadratic-weighted Cohen’s kappa of 0.564. Post hoc profiles varied across duration, visual quality, mechanical load and motor efficiency.

**Conclusions:** EXUS was technically feasible for synchronized IMU-video capture during FoCUS. The pilot supports multimodal acquisition data as a way to describe technical profiles and generate formative feedback hypotheses, but the post hoc indices are not validated competency measures.

## Introduction and Background

Point-of-care ultrasound (POCUS) and FoCUS are increasingly embedded in acute care and medical education because they provide immediate bedside information when performed by trained operators (1,2). Their usefulness, however, remains strongly operator dependent. The learner must integrate three-dimensional anatomy, two-dimensional ultrasound images, probe orientation, image optimization and clinical context while manipulating the transducer in real time (3,4).

FoCUS acquisition can be understood as a closed motor-visual loop. The operator moves the probe, observes the effect on the ultrasound image, modifies hand position, changes angle or pressure, and repeats this cycle until the desired cardiac window is obtained. This loop is partly cognitive, because the operator must recognize anatomy and decide whether the image is acceptable, but it is also motor and perceptual. A final saved image may therefore represent only the endpoint of a sequence of corrections, hesitations and stabilizing movements. For education, the pathway that produced the image may be as important as the image itself.

Competency tools for POCUS and cardiac POCUS have improved standardization of assessment, but many tools remain based on direct observation, checklists, global ratings or final image review (5,6). These approaches are essential, yet they provide limited information about the acquisition process that generated the image. For deliberate practice, learners benefit from specific feedback about what to improve and how their actions changed task performance (9-12).

This distinction is relevant because two operators may obtain similar final views through different technical strategies. One may reach the window quickly with stable movements and little image disruption, whereas another may require longer scanning, repeated high-amplitude adjustments or sustained mechanical load. Conversely, rapid acquisition is not necessarily good if it produces a poor-quality or unstable image. A process-oriented assessment should therefore be able to describe timing, motion economy and visual response together, rather than reducing the acquisition to a binary pass/fail decision or a single static image rating.

Motion-based ultrasound studies suggest that probe movement can differentiate novice and expert behavior, and that metrics such as angular velocity, frequency of movement or jerk may describe aspects of technical skill (7,8). A limitation of motion-only approaches is that they may not capture whether movement improved or degraded the ultrasound image. EXUS was designed to observe the same acquisition from two synchronized sources: a transducer-mounted IMU and the ultrasound video stream. The primary objective of this pilot study was to evaluate the technical feasibility of this IMU-video capture and fusion workflow during FoCUS. Secondary post hoc objectives were to describe exploratory operator profiles and candidate feedback indices.

## Materials and Methods

Study design and participants: This was a single-center observational pilot study of EXUS during FoCUS acquisition. Operators were pseudonymized as R1, R2, R3, FLW, E1 and E2 according to broad functional training level. To reduce indirect identifiability in this small single-center pilot, individual-level descriptors such as exact time of training, detailed clinical exposure and specific practice frequency are not reported at operator level. These characteristics were used only to inform interpretation of feasibility and exploratory acquisition profiles. Each technical session included two complete repetitions of the protocol; therefore, 6 technical sessions yielded 12 analytical sessions. Each analytical session included four FoCUS views, for 48 planned acquisitions. Operators and volunteers were adults able to provide informed consent. In addition, the six operators completed a separate 100-image Likert quality-rating task to estimate inter-rater agreement for still-frame ultrasound image quality assessment.

Ethics: The study was approved by the Comité Ético Científico de Ciencias de la Salud UC, Pontificia Universidad Católica de Chile (Protocol ID 260309007; Ordinary Session No. 05, April 9, 2026). All participants provided informed consent before participation.

EXUS acquisition and fusion: EXUS synchronized ultrasound video with IMU data obtained from the transducer. Data were fused in 1-second windows. IMU-derived features included gyroscope root-mean-square, acceleration root-mean-square, jerk, direction changes and stability-related measures. Video-derived heuristic features included image sharpness, frame-to-frame difference, temporal coherence and a composite visual quality estimate. These features were intended to describe acquisition behavior, not diagnosis. The technical configuration is summarized in Table 1, and Figure 1 illustrates the hardware and backend data-flow loop.

**Table 1.** EXUS technical configuration.

| Component | Specification | Role in EXUS | Data/output |
| --- | --- | --- | --- |
| Ultrasound system | Mindray M9; cardiac transducer; cardiac preset; video also stored internally by the ultrasound system | Clinical FoCUS image source | B-mode ultrasound video stream captured by EXUS |
| Video capture | AVerMedia GC551G2 capture device | Converted ultrasound video output into a computer-readable stream | MP4 video organized by attempt |
| IMU hardware | Initial IMU recorded as UNN4554-20948; replaced during the study by an MPU-6050; connected through a Raspberry Pi Pico controller with custom controller software | Measured transducer motion during acquisition | Gyroscope, acceleration, jerk, direction-change and stability-derived features |
| Backend computer | Lenovo ThinkPad; AMD Ryzen 7 Pro processor; 24 GB RAM; Ubuntu | Ran the EXUS_1 acquisition backend | Time-stamped acquisition events and synchronized data streams |
| EXUS_1 software | Python and OpenCV prototype; custom prototype reviewed by investigators | Captured video and IMU streams, recorded start/stop events and stored labels | Parquet data files, MP4 video and acquisition folders by attempt |
| Synchronization and quality control | Start/stop button in the user interface for each acquisition window; Likert entry in the UI; timing based on system clock; automatic IMU calibration before each attempt | Aligned protocol events, IMU and video for 1-second fused windows | Temporal coverage, fused-window count and IMU integrity checks; sensor failure detected by post-session data review |

**Figure 1.**
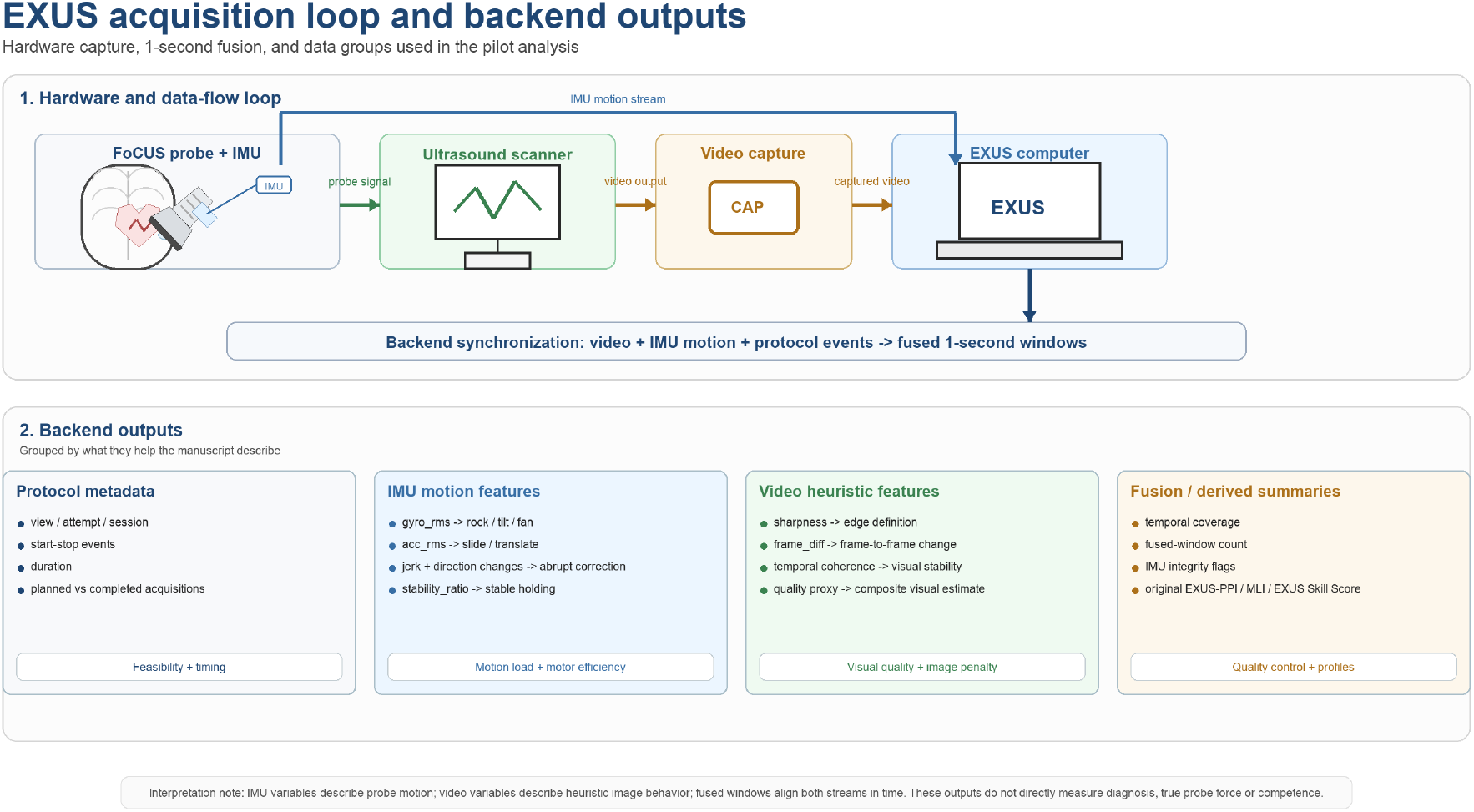
EXUS acquisition loop and backend outputs. The diagram summarizes how the transducer-mounted IMU, ultrasound video capture device and EXUS computer generated synchronized backend data for feasibility analysis and post hoc descriptive profiling. The backend outputs do not directly measure diagnosis, true probe force or operator competence.

EXUS hardware and software configuration: FoCUS was performed using a Mindray M9 ultrasound system with a cardiac transducer and cardiac preset. The ultrasound system stored video internally, but EXUS independently captured the video stream using an AVerMedia GC551G2 capture device. The backend ran on a Lenovo ThinkPad with an AMD Ryzen 7 Pro processor, 24 GB RAM and Ubuntu. EXUS_1 was implemented in Python with OpenCV. Start and stop events for each acquisition window, together with the operator Likert entry, were recorded through the user interface and time-stamped using the system clock. Output files were organized by attempt and included MP4 video and Parquet data files. The software captured IMU and video streams in real time and stored the data; it calculated the original EXUS-PPI during acquisition but did not provide real-time feedback. Other analyses were performed post hoc.

Primary feasibility endpoints: Feasibility was defined a priori as successful completion of the acquisition workflow. Endpoints included completion of analytical sessions, completion of planned acquisitions, start/stop events, video availability, fused IMU-video windows, temporal coverage at least 90%, complete human label entries and IMU integrity checks. The prespecified feasibility threshold was at least 80%. Wilson 95% confidence intervals were calculated for proportions.

Descriptive post hoc indices: The original EXUS-PPI was the system-exported pressure_proxy_index and is reported for traceability. It should not be interpreted as direct force or pressure. The exploratory post hoc indices were developed through theory-informed feature engineering and composite index construction. Raw IMU, video and timing variables were first mapped to clinically interpretable domains, including probe motion, image response, motion-video coupling, temporal efficiency and motor economy. Variables were then normalized and combined using explicit directional rules, so that higher MLI represented greater inferred mechanical load and higher EXUS Skill Score represented better descriptive technical performance. The MLI was defined post hoc as a descriptive mechanical load index combining IMU motion load, video-derived image penalty and motion-video coupling. Window-level variables were normalized using session percentiles p10-p90 and clipped to [0,1]. The EXUS Skill Score was a non-validated post hoc composite integrating duration, visual quality and IMU motor efficiency. These indices were not statistically trained, optimized or validated in this pilot sample and should be interpreted as transparent candidate constructs for future prospective validation. Rank-normalized 1-100 scales were used only for visualization within this sample. Additional operational detail, including the post hoc construct-development workflow and formulas, is provided in Supplementary Methods S1. The rationale for analyzing the best technically usable attempt was pragmatic and educational. This pilot was designed to determine whether EXUS could capture usable multimodal data and whether such data could describe operator profiles. When a known sensor failure occurred, treating the affected acquisition as operator behavior would have confounded technical feasibility with human performance. For this reason, post hoc profiles used the best technically usable attempt to describe what the system could observe when the acquisition was technically interpretable. This approach should be prespecified prospectively before any future validation or training-effect study.

Statistical analysis: Analyses were descriptive because of the pilot sample size. Operator profiles were summarized using medians or means as appropriate. Spearman correlation between normalized MLI and EXUS Skill Score was reported as exploratory only. No formal hypothesis testing of expertise classification was attempted. Inter-rater agreement for the 100-image Likert task was summarized using mean pairwise Cohen’s kappa, including weighted kappa because the Likert scale was ordinal. Use of generative AI tools is reported in the Declarations.

## Results

Technical feasibility: All 48 planned acquisitions were completed with video, start/stop events, fused IMU-video windows and complete human label entries. Temporal coverage was at least 90% in 47/48 acquisitions. IMU integrity endpoints also exceeded the 80% feasibility threshold, although a relevant IMU failure requiring sensor replacement and additional signal-integrity review was detected during one R2 attempt. Feasibility outcomes are summarized in Figure 2.

**Figure 2.**
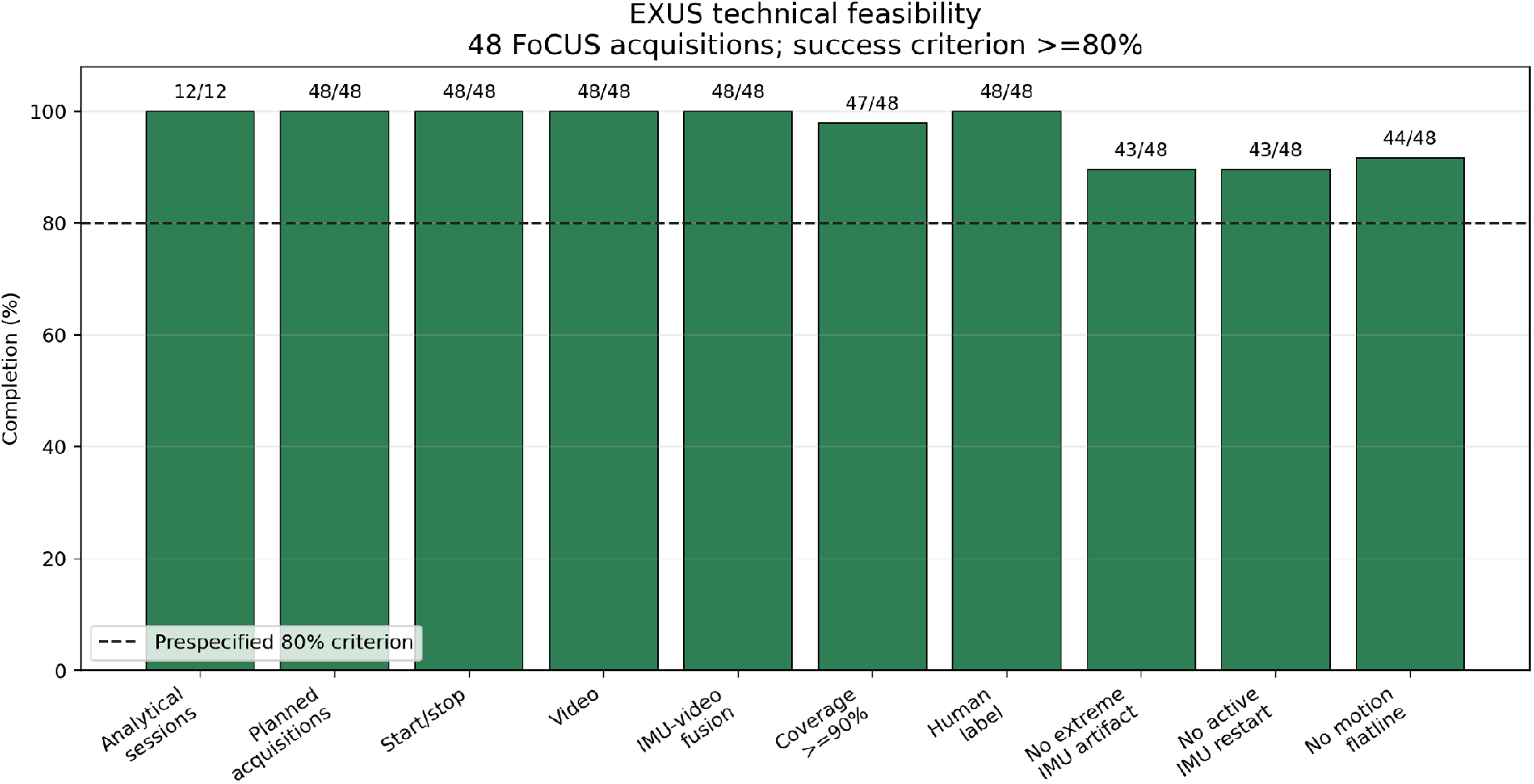
Technical feasibility endpoints. All evaluated endpoints met the prespecified 80% feasibility criterion. The figure summarizes proportions and denominators for the main capture and integrity checks.

Likert quality-rating agreement: For the separate 100-image Likert quality-rating task, the six final raters showed a mean pairwise exact agreement of 38.9%. Because the scale was ordinal, weighted agreement was more informative: mean pairwise linear-weighted Cohen’s kappa was 0.405 and mean quadratic-weighted Cohen’s kappa was 0.564, suggesting moderate agreement when one-point differences were accounted for.

Original EXUS-PPI and post hoc MLI: The original system-exported EXUS-PPI was retained as a traceability metric, but it did not provide a consistent description of the post hoc mechanical-load profile across operators. This supported development of the exploratory MLI, which more explicitly combined IMU motion, video-derived image degradation and motion-video coupling. This comparison should be interpreted as construct refinement rather than validation, because neither index measured true probe force.

Operator profiles: The best technically usable attempt per operator was used for descriptive profiling. R2 showed the longest acquisition duration and highest MLI. R1 was also relatively long but had lower mean mechanical load. R3 was fast with lower visual quality. FLW showed a balanced profile with high visual quality and IMU motor efficiency. E1 was fast and low-load but had lower IMU motor efficiency. E2 showed good visual quality with moderate-low mechanical load. These descriptive profiles are summarized in Table 2 and Figure 3.

**Table 2.** Operator technical profiles.

| Operator | Duration, s | Visual quality | MLI | IMU efficiency | Profile |
| --- | --- | --- | --- | --- | --- |
| R1 | 36.9 | 0.403 | 24.9 | -0.480 | Longer, low-load |
| R2 | 44.9 | 0.390 | 44.9 | -0.278 | Longest, highest-load |
| R3 | 8.3 | 0.271 | 26.7 | 0.532 | Fast, lower-quality |
| FLW | 11.5 | 0.420 | 26.2 | 0.583 | Balanced high-quality/efficient |
| E1 | 9.0 | 0.382 | 24.3 | -0.532 | Fast, low-load, lower IMU efficiency |
| E2 | 13.7 | 0.403 | 26.4 | 0.349 | Good quality, moderate-low load |

**Table 3.** Exploratory indices. Full operational formulas and normalization procedures are provided in Supplementary Methods S1.

| Index | Role in this manuscript | Inputs | Interpretation limits |
| --- | --- | --- | --- |
| Original EXUS-PPI | System-exported traceability metric | Motion stress, image resistance and micro-slip terms from fused windows | Not a direct force or pressure measurement |
| MLI | Post hoc descriptive mechanical load index | IMU motion load, video-derived image penalty and motion-video coupling | Not a validated pressure or competence score |
| EXUS Skill Score | Post hoc descriptive composite | Duration, visual quality and IMU motor efficiency | Not a validated competency or certification score |

**Figure 3.**
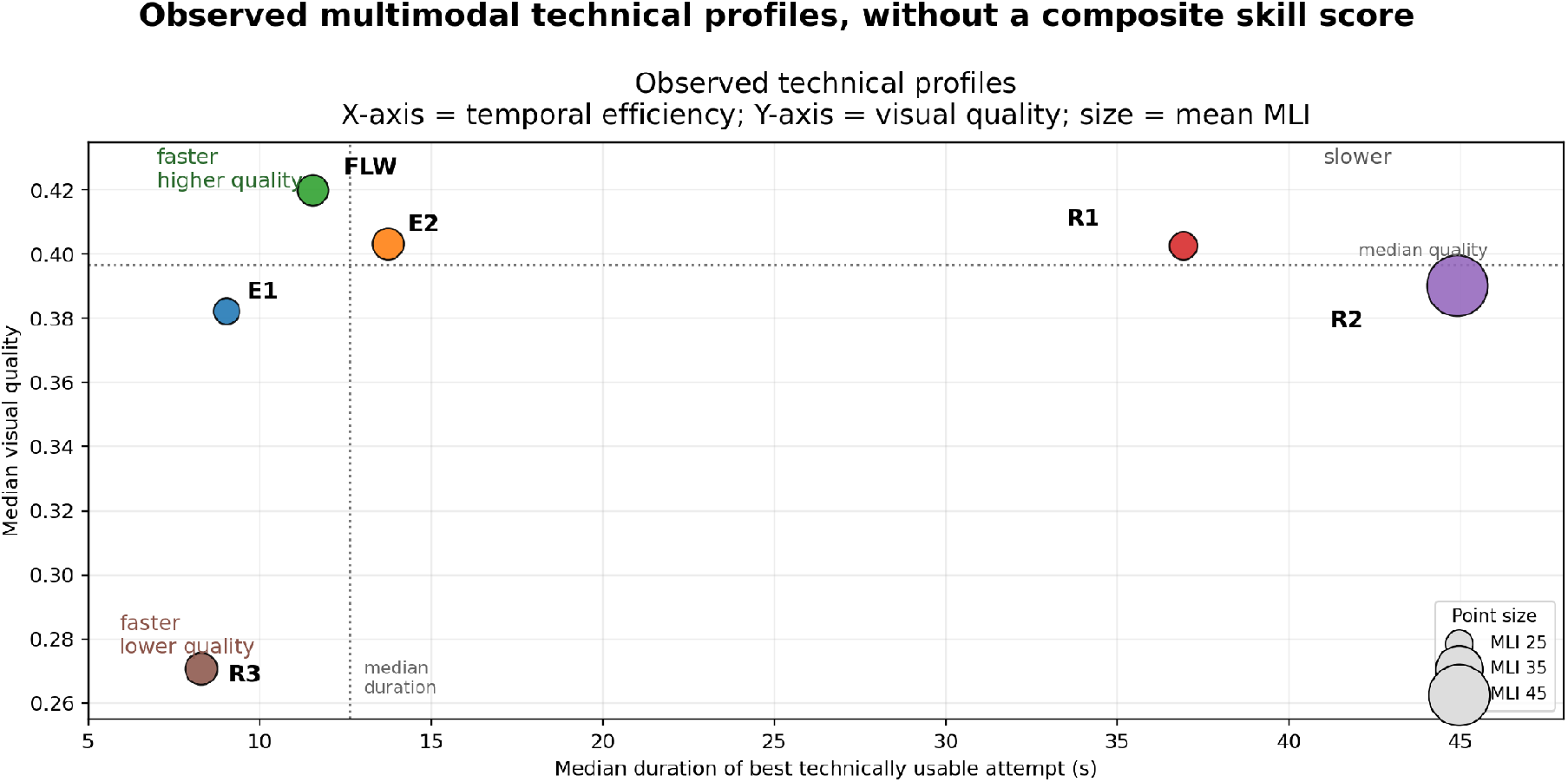
Observed technical profiles. The x-axis represents acquisition duration, the y-axis represents heuristic visual quality and point size represents mean MLI. The figure is descriptive and should be read with Table 2.

**Figure 4.**
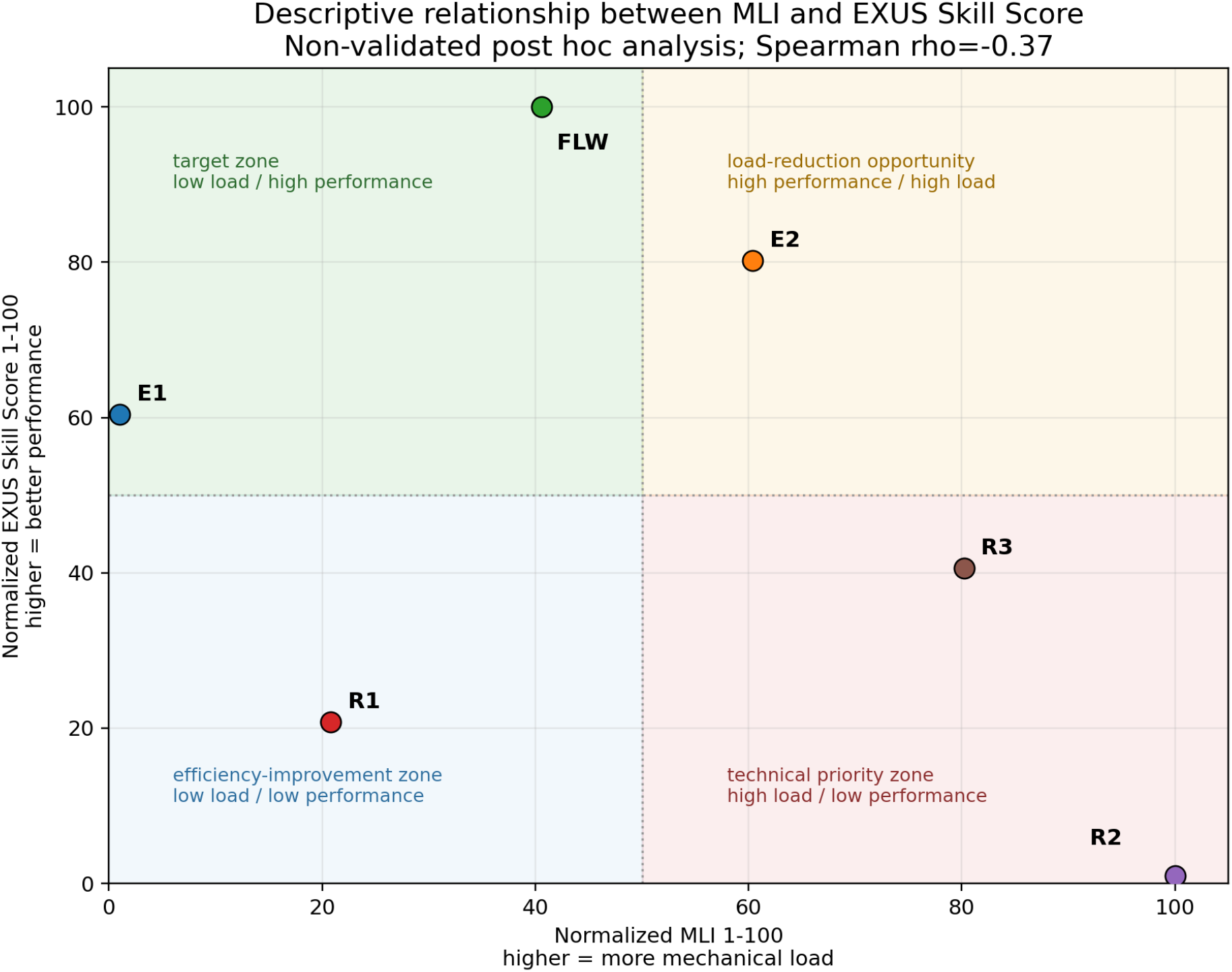
MLI and EXUS Skill Score. Both scores are derived from the same acquisition. MLI emphasizes mechanical load and adverse motion-video coupling, whereas the EXUS Skill Score emphasizes time, visual quality and motor efficiency. Quadrants are descriptive feedback zones.

Exploratory relationship of MLI and EXUS Skill Score: The normalized MLI-Skill Score plot suggested heterogeneous quadrants rather than a single expertise ranking. The Spearman rho was -0.37, indicating a weak-to-moderate inverse monotonic association in this small sample. This relationship should be interpreted as an exploratory visualization of two related but non-independent summaries, not as validation of either index.

## Discussion

This pilot study supports the technical feasibility of EXUS for synchronized IMU-video capture during FoCUS. The strongest finding is not validation of a competency metric, but successful acquisition, fusion and quality control of multimodal data in a real operator workflow. All planned acquisitions were completed, the key capture elements were available, and the predefined feasibility threshold was met across video, fusion, temporal coverage, human labels and IMU integrity endpoints. These results suggest that the acquisition process can be instrumented without interrupting the basic FoCUS workflow.

The observed IMU failure is also an important feasibility finding. In a study that proposes sensor-derived performance summaries, signal integrity is crucial for interpretation. A sensor restart, flatline or extreme artifact can mimic or obscure operator behavior. Detecting this failure during EXUS 1 supports the need for commercial-grade sensors, prospective signal-integrity rules and transparent handling of unusable acquisitions in future studies. Rather than invalidating the feasibility objective, this event defines one of the practical requirements for moving from prototype acquisition to educational or validation research.

The added value of combining IMU and video is that movement can be interpreted in relation to the visual result. IMU-only approaches can quantify acceleration, angular velocity or jerk, but cannot determine whether a movement improved the image, degraded it or represented efficient exploration. Video-only approaches can describe image quality but do not directly describe the physical actions that produced it. A large movement may be appropriate if it rapidly improves the window, whereas a small movement may be unhelpful if the image remains poor. EXUS links these two perspectives in time, allowing movement to be interpreted alongside sharpness, temporal coherence and frame-to-frame visual change.

This fusion is particularly relevant for formative feedback. Traditional feedback after FoCUS training often uses global language such as ‘improve probe handling’ or ‘optimize the image.’ Multimodal acquisition data could make feedback more specific: reduce high-load movements that do not improve the image, slow down during visually unstable segments, avoid repeated direction changes, or preserve a stable hand position once a satisfactory window appears. These are not conclusions from the current pilot, but examples of the type of feedback hypothesis that synchronized IMU-video data can generate.

The observed profiles are consistent with the educational concept that ultrasound performance is not a single dimension. Operators may be fast but visually unstable, slow but low-load, high-quality but mechanically inefficient, or balanced across domains. Such profiles may be more useful for formative feedback than a binary expert/non-expert label. This aligns with deliberate practice theory, where improvement depends on repeated task performance, informative feedback and targeted refinement of specific components of skill (9-12). In this framework, the purpose of EXUS is not to certify competence from a single acquisition, but to identify specific features of acquisition behavior that can be practiced deliberately.

The MLI and EXUS Skill Score should remain clearly framed as post hoc descriptive constructs. They are derived from overlapping sensor-video data and therefore cannot be treated as independent validation of each other. However, their weak-to-moderate inverse association in this pilot sample (Spearman rho = -0.37) suggests that they are not simply the same score expressed in opposite directions. Rather, they appear to capture partially related but non-equivalent aspects of acquisition behavior. MLI asks whether the acquisition showed mechanical load and adverse motion-video coupling, whereas the EXUS Skill Score asks whether the acquisition was temporally efficient, visually acceptable and motorically economical. This incomplete inverse relationship is useful because it allows operators to be described across four feedback-oriented zones: low load/high skill, high load/high skill, low load/low skill and high load/low skill. These zones should not be interpreted as validated categories, but as exploratory acquisition profiles that may help generate individualized deliberate-practice feedback in future studies. The 100-image Likert task adds a modest but useful quality-control element. Exact agreement across a 5-point scale was limited, as expected for ordinal image quality ratings, but weighted pairwise agreement was moderate when one-point differences were accounted for. This supports the feasibility of using human quality ratings as an exploratory reference signal, while also showing that still-frame Likert ratings should not be treated as a definitive validation standard. Future work should use prespecified rater training, blinded expert review and reliability targets if human image-quality assessment is used as an anchor for sensor-derived metrics.

A practical educational implementation would require translating these summaries into a small number of actionable feedback messages. For example, an operator with low visual quality and short duration may need to slow the search strategy and confirm anatomy, whereas an operator with good visual quality but high MLI may need to reduce unnecessary mechanical load once the window is obtained. In future iterations, this framework could evolve into a real-time feedback system in which transducer movement patterns are continuously interpreted together with automated ultrasound image-quality assessment, potentially using deep learning models trained for view recognition and image-quality scoring. Such a system could provide immediate formative cues during acquisition, helping learners adjust probe motion, stabilize useful windows, reduce non-productive mechanical load and repeat targeted tasks during deliberate practice.

Taken together, these findings support EXUS as a platform for feasibility-tested multimodal acquisition profiling and as a foundation for future real-time educational feedback. The next research step is not to claim that MLI or EXUS Skill Score measure competence, but to prospectively test whether predefined sensor-video summaries are reliable, interpretable and associated with external educational outcomes. These outcomes may include blinded expert ratings, longitudinal improvement during training, operator level, recent practice exposure and, eventually, response to real-time feedback supported by validated image-quality models.

## Limitations of the Study

This study included only 6 operators and 48 acquisitions, limiting statistical inference and external validity. The sample was intentionally small because the primary objective was feasibility. MLI and EXUS Skill Score were developed post hoc and are not validated competency measures. No direct force sensor was used, so MLI cannot be interpreted as true probe pressure. The video-derived quality variables were heuristic and should not be interpreted as machine-learning image-quality assessment. During the study, a relevant IMU device failure required sensor replacement and additional signal-integrity checks. This did not invalidate the feasibility objective, but it highlights the need for prospective sensor-integrity criteria and predefined handling rules for unusable attempts. No blinded expert panel independently rated all dynamic acquisitions for validation of technical performance. Although the 100-image Likert task showed moderate ordinal agreement, it remains an exploratory still-frame quality assessment with a small number of raters and does not fully represent dynamic acquisition quality. The best technically usable attempt was used for operator profiles to avoid interpreting known sensor failure as operator behavior; this approach is reasonable for feasibility profiling but should be prespecified prospectively. Finally, rank-normalized 1-100 displays are ordinal visualizations within this sample and should not be interpreted as absolute scores or thresholds. This preprint should therefore be read as a feasibility report, not as evidence that EXUS-derived indices are ready for clinical, educational or certification decisions.

## Conclusions and Future Research

EXUS was technically feasible for capturing and fusing transducer-mounted IMU and ultrasound video data during FoCUS. This pilot supports a multimodal framework for describing acquisition profiles and generating formative feedback hypotheses grounded in deliberate practice. The exploratory MLI and EXUS Skill Score should be treated as candidate descriptors for future study, not validated measures of pressure, performance or competence. Future studies should prospectively predefine these metrics, use robust commercial sensors, include direct force or pressure sensing when feasible, obtain blinded expert ratings and evaluate reliability across larger samples. The longer-term goal is to develop a real-time educational feedback system that integrates transducer movement patterns with automated ultrasound view recognition and image-quality assessment, potentially using validated deep learning models. Such a system could help learners adjust probe motion during acquisition, reduce non-productive mechanical load, stabilize useful windows and repeat targeted tasks as part of deliberate FoCUS practice.

## Declarations

### Ethics approval and consent to participate

Ethics approval and consent to participate: This study involved human participants and human-derived ultrasound acquisition data. Ethics approval was obtained from the Comité Ético Científico de Ciencias de la Salud UC, Pontificia Universidad Católica de Chile (Protocol ID 260309007; Ordinary Session No. 05, April 9, 2026). All participants provided informed consent before participation.

### Consent for publication

Participants provided consent for use of anonymized study data for scientific publication. No identifiable individual images or videos are included in this manuscript.

### Clinical trial registration

Clinical trial registration: Not applicable; this was an observational feasibility pilot and did not prospectively assign participants to a clinical or educational intervention.

### Availability of data and materials

The datasets generated and/or analysed during the current study are not publicly available due to participant privacy and institutional restrictions but are available from the corresponding author on reasonable request, subject to ethics and institutional approval.

### Authors’ contributions

FR, DA and BL conceived the study. FR developed the EXUS acquisition workflow. FR collected the data. FR analyzed and interpreted the data. FR drafted the manuscript. All authors critically revised the manuscript and approved the final version.

### Use of artificial intelligence tools

Generative AI-assisted tools, including OpenAI ChatGPT/Codex, were used to support software prototyping, code debugging, data-processing script drafting, exploratory visualization formatting, manuscript language editing and preparation of explanatory text.

All AI-assisted outputs were reviewed and modified by the authors. The study design, ethical conduct, outcome reporting, final analytic choices, result interpretation, reference verification and final manuscript content were determined and approved by the human investigators. No AI tool was listed as an author. No identifiable participant information or raw ultrasound videos were intentionally submitted to external generative AI platforms. The authors remain fully accountable for the accuracy, integrity and originality of the work.

## Abbreviation

EXUS: Expert in Ultrasound Skills
FoCUS: focused cardiac ultrasound
IMU: inertial measurement unit
MLI: Mechanical Load Index
POCUS: point-of-care ultrasound
PPI: pressure proxy index

## Acknowledgements

The authors thank the operators and volunteers who participated in the pilot study. The authors also thank the Emergency Medicine Section of the Department of Internal Medicine, Pontificia Universidad Católica de Chile, and Hospital Clínico UC Christus for supporting the clinical and educational setting in which this feasibility study was conducted.

