## Supplementary material for "Expert in Ultrasound Skills: Feasibility of an IMU-video platform to describe technical profiles during focused cardiac ultrasound. Pilot study": Suplement 1 (from another version of this preprint: version 1, 2026-06-18)

### Supplementary Methods S1

*Plain-language operational definitions of EXUS-derived exploratory indices*

> **Purpose of this supplement**
> This appendix is written to make the EXUS-derived indices understandable and reproducible. The indices are post hoc descriptors from a feasibility pilot. They should not be read as validated statistical models, direct pressure measurements or competency scores.

### How to Read the Indices

The EXUS_1 backend captured synchronized ultrasound video and probe-mounted IMU signals. These raw signals were fused into 1-second windows. The indices below summarize those windows from different angles: traceability, inferred mechanical load and descriptive technical performance.

A useful way to read this supplement is to imagine the acquisition as a short clinical procedure that leaves a digital trace. The button presses define when the acquisition starts and ends. The IMU describes how the probe moved in the operator's hand. The video describes how the ultrasound image changed over time. EXUS then compresses those traces into window-level variables before building any index.

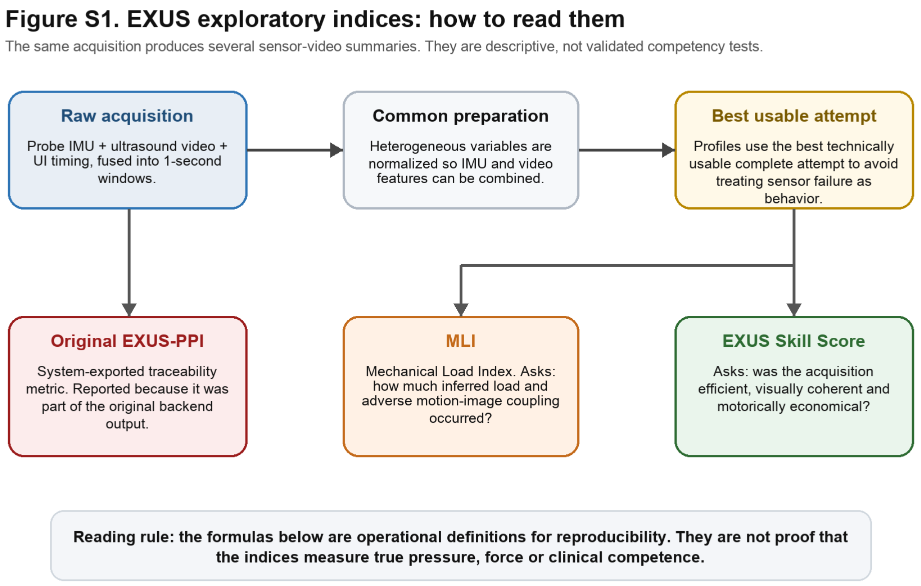


*Figure S1. The original EXUS-PPI is retained for traceability. MLI and EXUS Skill Score are post hoc descriptive summaries of the same acquisition, oriented to different questions.*

### How the Post Hoc Indices Were Built

The post hoc analysis followed a transparent construct-development workflow rather than a predictive modelling workflow. No statistical model was trained, and no automatic weight optimization was performed in this six-operator dataset. The aim was to translate clinically plausible acquisition behavior into reproducible exploratory descriptors.

> **Methodological label**
> A concise name for this process is theory-informed feature engineering and composite index construction. Another acceptable phrasing is exploratory operationalization of sensor-derived constructs.

**Table S1. Post hoc construct-development workflow**

| Layer | Decision | EXUS application |
|---|---|---|
| Clinical construct | Define the question before the formula | MLI asks whether motion coincided with adverse image response. EXUS Skill Score asks whether the acquisition was efficient, coherent and motorically economical. |
| Observable domains | Map the construct to signals | Probe movement from IMU, image behavior from video and timing from UI start/stop. |
| Feature engineering | Convert raw signals to variables | Examples include gyro_rms, acc_rms, jerk_mean, stability_ratio, sharpness_mean, frame_diff_mean and quality_mean. |
| Scaling and direction | Put variables on comparable scales | Variables were normalized and oriented so higher MLI means more inferred load, while higher EXUS Skill Score means a better descriptive profile. |
| Composite construction | Combine domains with explicit weights | Weights were theory-informed operational choices, not weights learned from the pilot dataset. |
| Plausibility check | Look for coherent descriptive behavior | The indices were compared with operator profiles and with each other as hypothesis-generating summaries. |

This distinction is important. The indices are not black-box predictions, but they are also not validated measurements. They are explicit operational definitions that can be inspected, criticized, reproduced and prospectively tested in EXUS 2.

### From Raw Signals to Backend Variables

For a clinical reader, the most important point is that EXUS does not start with abstract scores. It first records simple signals during acquisition: time from the user interface, movement from the probe-mounted IMU and frame-level behavior from the ultrasound video. These signals are summarized every 1 second before any exploratory index is calculated.

> **Clinical reading**
> The variables below are engineering summaries of acquisition behavior. They describe how the probe moved and how the video behaved during the scan. They are not direct measurements of pressure, diagnostic image quality or operator competence.

**Table S2. How raw EXUS signals became window-level variables**

| Raw source | Backend operation | Output variable | Clinical meaning |
|---|---|---|---|
| UI start/stop | Time between acquisition start and stop | elapsed_sec | How long the operator needed to complete the acquisition. |
| Gyroscope x/y/z | Root mean square of calibrated rotational velocity samples | gyro_rms | How much the probe rotated, rocked or tilted. |
| Accelerometer x/y/z | Root mean square of calibrated acceleration samples | acc_rms | How much translational acceleration occurred. |
| Consecutive acceleration samples | Mean magnitude of acceleration change over time | jerk_mean | How abrupt or jerky the movement was. |
| IMU signal | Proportion of samples below a stability threshold | stability_ratio | How much of the window looked mechanically stable. |
| Video frame | Variance of the Laplacian in grayscale | sharpness_mean | A heuristic estimate of image sharpness. |
| Consecutive video frames | Mean absolute grayscale difference between frames | frame_diff_mean | How much the image changed from frame to frame. |
| Video + IMU | Temporal video coherence combined with IMU stability | quality_mean | A heuristic summary of visual coherence and stability. |

For example, if an operator rocks the probe repeatedly, gyro_rms tends to increase. If the operator makes abrupt corrections, jerk_mean tends to increase. If the ultrasound image changes substantially from one frame to the next, frame_diff_mean tends to increase. These variables therefore describe observable aspects of acquisition behavior, but they do not by themselves say whether the scan was clinically adequate.

### Variable Map

Table S3 translates the same variables into their role in the exploratory indices. This table is meant as a reader's map before the formulas appear.

**Table S3. Backend variables and their role in the exploratory indices**

| Source | Variable | Plain-language meaning | Used mainly for |
|---|---|---|---|
| UI clock | elapsed_sec | Time needed to complete a technically usable acquisition. | EXUS Skill Score |
| IMU | gyro_rms | Rotational probe movement, such as rocking or tilting. | MLI; motor efficiency |
| IMU | acc_rms | Translational acceleration of the probe. | MLI |
| IMU | jerk_mean | Abrupt changes in probe movement. | MLI; motor efficiency |
| IMU | stability_ratio | Proportion of the window with stable IMU behavior. | Original EXUS-PPI; visual heuristic |
| Video | sharpness_mean | Heuristic image sharpness based on video texture. | Original EXUS-PPI; MLI |
| Video | frame_diff_mean | Frame-to-frame image change; higher values suggest more visual instability. | Original EXUS-PPI; MLI |
| Video + IMU | quality_mean | Heuristic combination of temporal image coherence and IMU stability. | Original EXUS-PPI; EXUS Skill Score |

### Normalization: Why the Variables Are Rescaled

The backend variables have different units and ranges. For example, IMU rotation, acceleration, jerk, image sharpness and frame-to-frame image change cannot be added directly in their raw units. A robust percentile-based normalization placed them on a common 0-1 scale before weighting.

Clinically, this is similar to converting different vital signs or laboratory values into a common reference scale before combining them. The procedure does not make the variables equivalent; it only prevents one variable from dominating the index simply because it has larger numerical units.

> **Robust within-session normalization**
> n(x) = clip((x - p10) / (p90 - p10), 0, 1)
>
> p10 and p90 = session-specific 10th and 90th percentiles
> clip(..., 0, 1) = values below 0 are set to 0; values above 1 are set to 1

This is a pragmatic robust scaling approach. It reduces the influence of isolated spikes, but the p10-p90 range should be prespecified and sensitivity-tested in future validation studies.

### Original EXUS-PPI: Traceability Metric

The original EXUS-PPI was the system-exported pressure_proxy_index generated by the EXUS_1 backend. It is reported because it was part of the original software output. It should not be interpreted as measured probe pressure.

> **Plain-language reading**
> The original EXUS-PPI combines three intermediate ideas: motion stress, image resistance and micro-slip. These components are operational software terms, not validated physical measurements.

In clinical terms, the original EXUS-PPI can be read as an early software attempt to summarize whether movement and image behavior looked unfavorable during a 1-second window. It remains useful for auditability because it was exported by the original system, but the current manuscript does not treat it as a validated pressure or force measurement.

**Original EXUS-PPI computational definition**

frame_diff_norm = n(frame_diff_mean)
temporal_coherence = 1 - frame_diff_norm
quality_mean = 0.60 * temporal_coherence + 0.40 * stability_ratio

motion_stress = 0.50 * n(jerk_mean) + 0.40 * n(gyro_rms) + 0.10 * n(acc_rms)
image_good = 0.60 * quality_mean + 0.40 * n(sharpness_mean)
image_resistance = 1 - image_good
micro_slip = n(jerk_mean) * frame_diff_norm

Original EXUS-PPI = 100 * (0.50 * motion_stress + 0.35 * image_resistance + 0.15 * micro_slip)

### Mechanical Load Index

The Mechanical Load Index (MLI) was developed post hoc to describe inferred mechanical load and adverse motion-image coupling. It is easier to read as a layered algorithm than as a single formula.

The clinical intuition behind MLI is that movement is not always bad. A learner may need to move the probe while searching for a window. MLI gives more importance to movement when it appears together with image deterioration or instability. In other words, it tries to describe the part of movement that may represent unnecessary mechanical load or inefficient coupling between hand motion and image response.

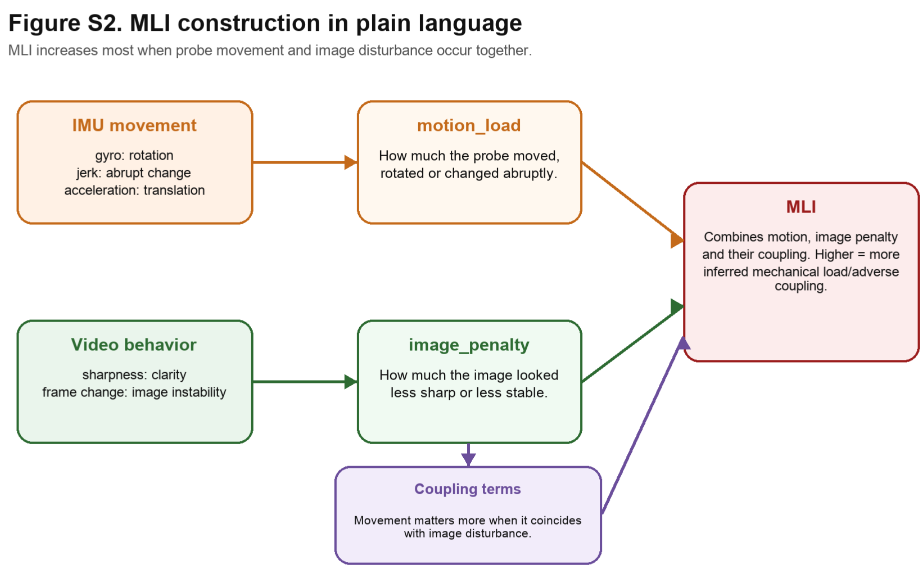


*Figure S2. MLI increases when movement-derived load and video-derived image penalty occur together. Movement alone is not automatically interpreted as poor performance.*

#### *MLI in Four Steps*

- Step 1: quantify probe movement from IMU rotation, abrupt movement and acceleration.
- Step 2: quantify image penalty from reduced sharpness and reduced temporal coherence.
- Step 3: increase the value when movement and image penalty occur in the same window.
- Step 4: average window-level MLI within each view, then average the four view-level means for the operator profile.

**Why average instead of using p75 or p90?**
The final manuscript version uses the mean of view-level means. This avoids selecting an arbitrary high percentile and gives each echocardiographic view equal contribution in the best technically usable attempt.

This aggregation was chosen for transparency. The aim was to describe the overall mechanical-load profile of a usable attempt, not to select the worst seconds of the acquisition. Future studies can test whether mean, p75 or other summaries are more reliable, but those choices should be prespecified.

> **MLI computational definition**
> motion_load = 0.45 * n(gyro_rms) + 0.35 * n(jerk_mean) + 0.20 * n(acc_rms)
> temporal_coherence = 1 - n(frame_diff_mean)
> image_penalty = 1 - (0.55 * n(sharpness_mean) + 0.45 * temporal_coherence)
>
> coupled_load = motion_load * image_penalty
> dynamic_damage = motion_load * n(frame_diff_mean) * image_penalty
>
> MLI_window = 100 * (0.30 * motion_load + 0.25 * image_penalty
> + 0.20 * coupled_load + 0.25 * dynamic_damage)
>
> Operator-level MLI = mean of four view-level means from the best technically usable attempt

MLI excludes acquisition duration, Likert ratings, number of windows, the original EXUS-PPI, the EXUS Skill Score and external expert ratings. Higher MLI values indicate greater inferred mechanical load and/or adverse motion-video coupling within this dataset only.

### EXUS Skill Score

The EXUS Skill Score was developed post hoc as a descriptive technical-performance composite. It asks a different question from MLI: was the acquisition efficient, visually coherent and motorically economical?

This score should be read as a compact acquisition profile, not as a grade. A higher value means that, within this pilot dataset, the completed acquisition was relatively shorter, visually more coherent and associated with more economical IMU movement. It does not prove expertise and it does not replace expert judgement.

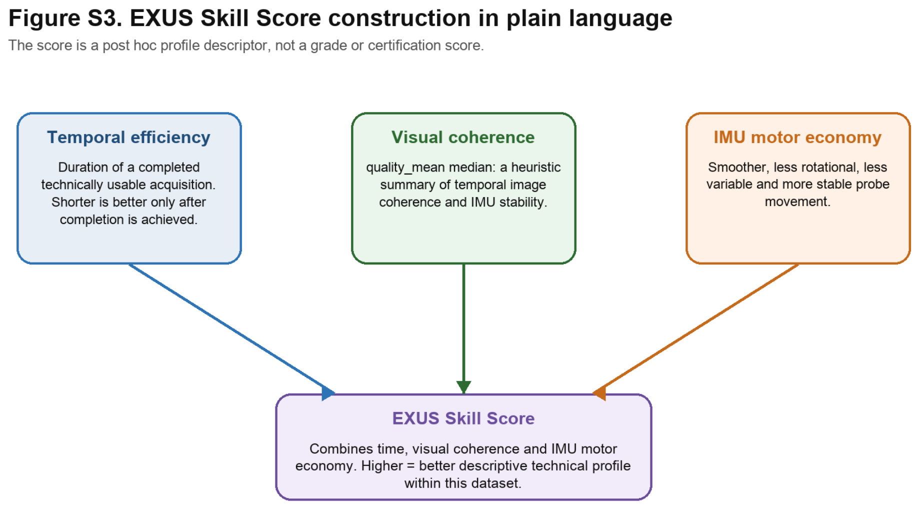


*Figure S3. EXUS Skill Score combines three domains. It does not include MLI, original EXUS-PPI, Likert ratings or number of windows.*

#### *The Three Domains*

| Domain | What it means | Direction |
|---|---|---|
| Temporal efficiency | Duration of a completed technically usable acquisition. | Shorter is better only after completion is achieved. |
| Visual coherence | Median quality_mean across 1-second windows. | Higher is better. |
| IMU motor economy | Smoother, less rotational, less variable and more stable movement. | Higher is better. |

> **How to read the IMU motor economy term**
> This is not a biomechanical model. It is a compact post hoc descriptor that rewards smoother and more consistent probe movement and penalizes abrupt, rotational or highly variable movement.

The IMU component is intentionally simple. It penalizes typical abruptness, typical rotation and variability in abruptness. It also includes a small consistency term: whether the IMU signal stayed near a common movement pattern for much of the acquisition.

#### *Two Technical Terms*

| Term | Reader-friendly meaning |
|---|---|
| log(1 + x) | A transformation that compresses isolated high IMU values so a single spike does not dominate the score. The '+1' allows zero values to be transformed. |
| IQR | Interquartile range, calculated as p75 - p25. It describes central variability. A higher jerk IQR means movement abruptness varied more during the acquisition. |
| IMU modal stability ratio | A consistency term. If most IMU values stay near the most common movement range, the ratio is higher. If movement is scattered across many ranges, the ratio is lower. |

> **EXUS Skill Score computational definition**
> Visual coherence component:
> frame_diff_norm = n(frame_diff_mean)
> temporal_coherence = 1 - frame_diff_norm
> quality_mean = 0.60 * temporal_coherence + 0.40 * stability_ratio
> quality_mean_median = median quality_mean across windows in the acquisition
>
> IMU motor economy component:
> log_jerk_median = log(1 + jerk_mean_median)
> log_gyro_median = log(1 + gyro_rms_median)
> log_jerk_iqr = log(1 + jerk_mean_iqr)
>
> IMU_motor_efficiency = -0.35 * z(log_jerk_median)
> -0.30 * z(log_gyro_median)
> -0.20 * z(log_jerk_iqr)
> +0.15 * z(imu_modal_stability_ratio)
>
> EXUS Skill Score = -0.40 * z(elapsed_sec)
> +0.35 * z(quality_mean_median)
> +0.25 * z(IMU_motor_efficiency)

Here z() denotes standard z-score normalization within the pilot analysis dataset. The EXUS Skill Score excludes MLI, original EXUS-PPI, explicit mechanical-load terms, Likert ratings, number of windows and external expert ratings.

### Interpretation Boundaries

The indices are useful only if their boundaries are explicit. Table S4 summarizes what each index can and cannot support in the current feasibility pilot.

**Table S4. Interpretation boundaries for EXUS-derived exploratory indices**

| Index | Question asked | Higher value means | Do not interpret as |
|---|---|---|---|
| Original EXUS-PPI | What did the original backend export? | More motion stress, image resistance and micro-slip. | Measured pressure or validated mechanical load. |
| MLI | How much inferred load and adverse motion-video coupling occurred? | More inferred mechanical load/ coupling. | True force, pressure or competence. |
| EXUS Skill Score | Was the acquisition efficient, visually coherent and motorically economical? | Better descriptive technical profile. | Certification, grading or independent validation. |

### Rank-normalized 1-100 Figures

Some figures display MLI and EXUS Skill Score on a rank-normalized 1-100 scale to improve visual readability in a six-operator dataset. This is an ordinal display only.

> **Visualization-only scaling**
> score_1_to_100 = 1 + 99 * (rank - 1) / (N - 1)
>
> N = number of operators
> For MLI: higher = more mechanical load
> For EXUS Skill Score: higher = better descriptive technical profile

> **Final caution**
> The 1-100 displays are within-sample visualizations. They are not absolute scores, validated thresholds or externally generalizable cutoffs.
